# An Alzheimer’s disease pathway uncovered by functional omics: the risk gene *CELF1* regulates *KLC1* splice variant E expression, which drives Aβ pathology

**DOI:** 10.1101/2022.02.28.22271320

**Authors:** Masataka Kikuchi, Justine Viet, Kenichi Nagata, Masahiro Sato, Géraldine David, Yann Audic, Michael A. Silverman, Mitsuko Yamamoto, Hiroyasu Akatsu, Yoshio Hashizume, Kyoko Chiba, Shuko Takeda, Shoshin Akamine, Tesshin Miyamoto, Ryota Uozumi, Shiho Gotoh, Kohji Mori, Manabu Ikeda, Luc Paillard, Takashi Morihara

## Abstract

In an era when numerous disease-associated genes have been identified, determining the molecular mechanisms of complex diseases is still difficult. The *CELF1* region was identified by genome-wide association studies as an Alzheimer’s disease (AD) risk locus. Using transcriptomics and cross-linking and immunoprecipitation sequencing (CLIP-seq), we found that CELF1, an RNA-binding protein, binds to *KLC1* RNA and regulates its splicing. Analysis of two brain banks revealed that *CELF1* expression is correlated with inclusion of *KLC1* exons downstream of the CELF1-binding region identified by CLIP-seq. In AD, low *CELF1* levels result in high levels of *KLC1* splice variant E (*KLC1_vE*), an amyloid-β (Aβ) pathology-driving gene product. Cell culture experiments confirmed regulation of *KLC1_vE* by CELF1. Analysis of mouse strains with different propensities for Aβ accumulation confirmed that *Klc1_vE* drives Aβ pathology. Using omics methods, we revealed the molecular pathway of a complex disease supported by human and mouse genetics.

## Introduction

The number of disease-associated loci that have been identified by powerful genome-wide association studies (GWASs) is rapidly increasing, but functional analysis of the identified loci and studies of how the risk genes contribute to diseases are still challenging. As a result, there has been precipitous increase in the number of associated loci and genes identified whose function in disease is unknown^1^. Many researchers have suggested that the innovative use of functional omics will be crucial to interpret GWAS results and reveal molecular pathways in complex diseases in the post-GWAS era, but concrete strategies are still being explored^1,2,3,4^.

In Alzheimer’s disease (AD), there is a sharp disparity between the criticality of the disease, including the large number of patients and the burden of care, and the availability of treatments. There is also a large gap between the complexity of AD and the paucity of molecular pathogenic mechanisms known to researchers. As a result, the number of target molecules for new drug development is extremely limited^5^. Therefore, there is a strong need to identify new molecular pathways for AD that are supported by genetics, with the added reason that drugs that target genetically supported molecular pathways have a higher success rate in clinical trials. *CELF1*, studied here, has been identified as a risk locus in four large GWASs^6,7,8,9^, but similar to many other risk loci, the lead single nucleotide polymorphisms (SNPs) differ among genetic studies, and all of these SNPs are in noncoding regions. Therefore, as with many other loci, it is difficult to analyze the function of the *CELF1* locus using genomic variation information.

Previously, we identified *KLC1* splice variant E (*KLC1_vE*) as a gene product that drives amyloid-β (Aβ) pathology, a core pathology of AD, by performing an omics study that differed from a GWAS^10^. We noted that the DBA/2 mouse strain accumulates less Aβ than the C57BL/6 and SJL strains and analyzed the expression profiles of genes specific to DBA/2 mice. We found that the *Klc1* allele of DBA/2 mice reduced Aβ accumulation by decreasing *Klc1_vE* expression. In cultured cells, we confirmed that knockdown of *KLC1_vE* suppressed Aβ production. In the human brain, we also found that the *KLC1_vE* level was higher in AD patients than that in control subjects, although the regulatory mechanism of *KLC1_vE* in the human brain remains to be elucidated. Our unique transcriptomics approach focusing on mouse genetic backgrounds revealed that aberrant splicing of *KLC1* drives Aβ pathology.

The goal of this study was to elucidate the function of the *CELF1* locus and to uncover new molecular pathways for the development of AD. The impetus for this study originated from the integration of two omics approaches: exon array analysis indicated that *CELF1* knockdown resulted in highly altered exon expression of *KLC1*, while cross-linking immunoprecipitation sequencing (CLIP-seq) revealed binding of the CELF1 protein to *KLC1* RNA. Two types of analyses, reverse transcription quantitative polymerase chain reaction (RT-qPCR) and RNA sequencing (RNA-seq), of two brain banks, including one with Japanese and one with American Caucasian (AMP-AD Mayo^11^) donors, revealed that *CELF1* expression levels are strongly correlated with *KLC1* splicing, including *KLC1_vE*. Cell culture experiments confirmed regulation of *KLC1_vE* by *CELF1*. Analysis of seven mouse strains reported to have different propensities for Aβ pathology reconfirmed that the *Klc1* allele and *Klc1_vE* expression levels determine the propensity for development of Aβ pathology. By effectively utilizing such a wide variety of omics approaches, we discovered that decreased expression of the risk gene *CELF1* upregulates *KLC1_vE* expression and promotes Aβ pathology, which is a new molecular pathway for the development of AD.

## Results

### Integration of two omics approaches unexpectedly uncovered a strong link between CELF1 and KLC1

CELF1 is an RNA-binding protein that regulates the expression and splicing of many genes. CELF1 plays a role in cardiac development, myotonic dystrophy, and cancer^12,13^, but its function in AD has not been elucidated, even though the *CELF1* locus has been associated with AD^6,7,8,9^. We found a direct relationship between CELF1 and *KLC1* in data obtained by integrating two omics studies that we previously conducted^14,15^. First, transcriptomics data from HeLa cells showed that three of the ten exons most strongly altered by *CELF1* knockdown were *KLC1* exons (Fig. 1a). Second, CLIP-seq, which comprehensively identifies RNA regions that are bound by RNA-binding proteins, revealed that the CELF1 protein binds to *KLC1* RNA in HeLa cells (Fig. 1b). Specifically, a major CELF1 binding cluster (Fig. 1b) was detected on a complex alternative exon of *KLC1* labeled as exon 15 in previous studies^16,17^. These findings were confirmed by detection of “cross-linking-induced mutation sites”, which revealed the positions of nucleotides within exon 15 that interacted with CELF1 protein in CLIP-seq experiments^18^ (Fig. 1c). These results indicate that the regulation of *KLC1* exon expression by CELF1 observed in Fig. 1a is likely a direct effect of CELF1 binding to *KLC1* RNA. These two omics approaches suggest that CELF1 directly and potently regulates *KLC1*. Because *CELF1* is located in a risk locus for AD^6,7,8,9^ and *KLC1* regulates amyloid pathology^10,19^, the core pathology of AD, we considered whether this unexpected discovery might lead to a new understanding of AD pathogenesis in humans.

**Fig. 1.**
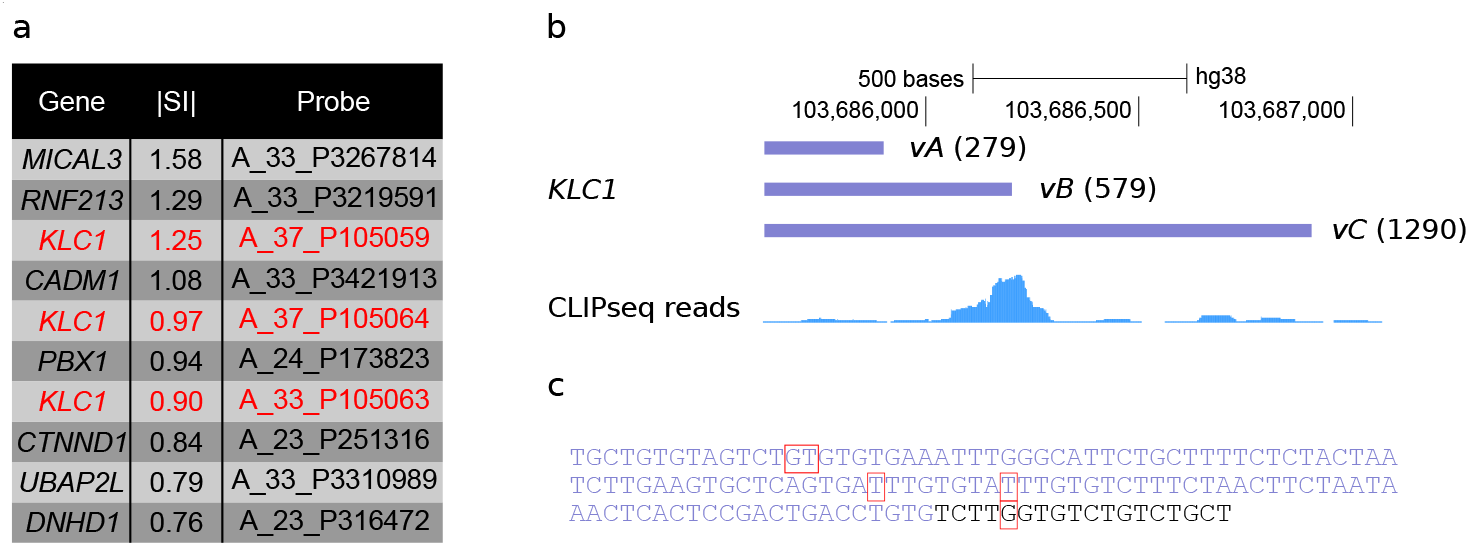
Regulation of *KLC1* splicing by CELF1 in HeLa cells demonstrated by exon array and cross-linking and immunoprecipitation sequencing (CLIP-seq) **a**. The top ten SurePrint G3 human exon array probes (Agilent) with the highest splicing indices (in absolute values) following *CELF1* knockdown. The splicing index (SI) is the log2 ratio of the normalized probe intensity in depleted cells to the probe intensity in control cells. Three probes for the *KLC1* gene are shown in red. **b**. Binding of CELF1 protein to *KLC1* RNA revealed by CLIP-seq. The genomic location of alternative exon 15, which is present in splice variants A, B, and C of *KLC1* RNA (upper), is shown. The CELF1 binding site in exon 15 of *KLC1* was revealed by the accumulation of CLIP-seq reads (bottom). Please note the accumulation of reads at the boundary between *vB* exon 15 and *vC* exon 15. **c**. Sequence of the 3’ end of *vB* exon 15 (purple nucleotides) and the downstream region (black nucleotides, specific for *vC* exon 15). Cross-link induced mutation sites in CLIP-seq are shown in red boxes.

### In the human brain, *CELF1* expression levels correlate with those of the major splice variants of *KLC1*, including *KLC1_vE*

To examine whether the regulation of *KLC1* by *CELF1* observed in cultured cells also occurs in the human brain, we quantified the expression levels of *CELF1* and the major splice variants of *KLC1* in autopsied brains. A hypothesis-free study identified *KLC1_vE* as a gene product that drives Aβ pathology in AD^10,19^. In brains of human AD patients, *KLC_vE* levels are elevated, but the molecular mechanisms that regulate *KLC1_vE* expression in the human brain remain elusive. Thus, the unexpected finding of functional omics analysis that CELF1 regulates expression of *KLC1* exons (Fig. 1) prompted us to investigate this issue.

To determine whether *CELF1* and *KLC1* coexist in the same cells in the brain, which is a prerequisite for the binding to and regulation of *KLC1* by CELF1 observed in cultured cells, we examined a single-cell RNA sequencing database^20^ (PanglaoDB, https://panglaodb.se/), which demonstrated that *CELF1* and *KLC1* coexist in the same cells, mainly consisting of neurons (Supplementary Fig. 1a).

We then measured the levels of *CELF1* and major *KLC1* variants in AD (n=10) and control (n=14) brains from Japanese individuals. We found a trend toward decreased *CELF1* expression (−24.4%, p=0.090) in AD brains compared with that in control brains (Fig, 2a). The decrease in the *CELF1* level was significant (–31.9%, p=0.021) when samples were differentiated by risk SNPs (rs10838725)^8^ for *CELF1* instead of pathological diagnosis (Fig. 2b). Furthermore, we found a strong correlation between the expression levels of *CELF1* and those of most major splice variants of *KLC1. KLC1_vA* (R^2^=0.21, p=0.023) tended to be positively correlated with *CELF1*, and *KLC1_vB* (R^2^=0.44, p=0.0004) was significantly positively correlated with *CELF1* (Fig. 2c). *KLC1_vC* (R^2^=0.61, p<0.0001), *vD* (R^2^=0.38, p=0.001), and *vE* (R^2^=0.64, p<0.0001) were significantly inversely correlated with *CELF1*. Next, to examine whether secondary effects of AD pathology are involved in these strong correlations between *CELF1* and *KLC1* splice variants, we analyzed only control brains without AD pathology. Although the statistical significance was reduced because of the lower number of samples, the correlations between *CELF1* and *KLC1* splice variants were also observed in control brains (Supplementary Fig. 2). Taken together, these observations suggested that the relationship between CELF1 and *KLC1* splice variants, including *KLC1_vE*, may be a direct and fundamental relationship in the brain and not a consequence of AD pathology.

**Fig. 2.**
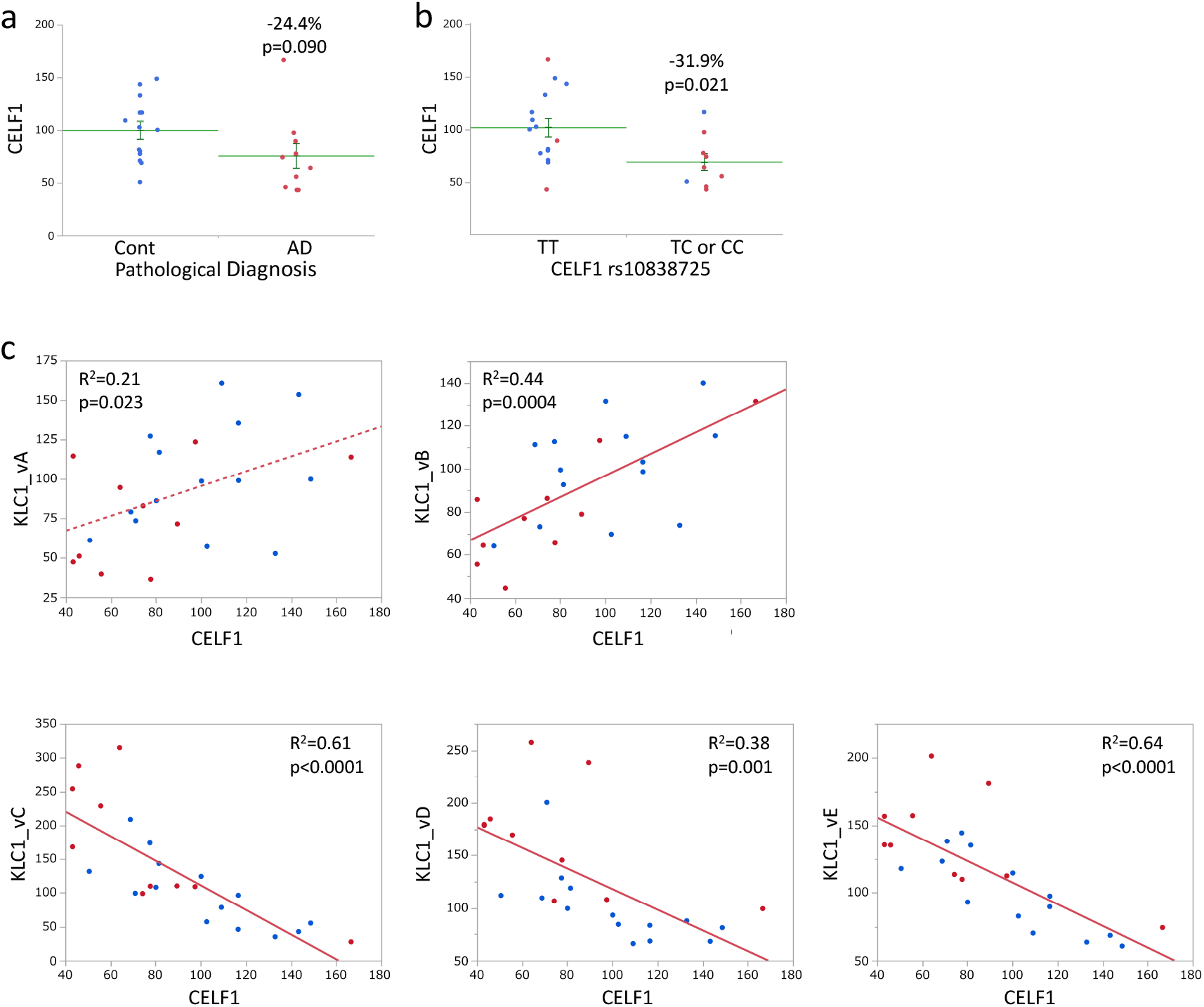
Expression levels of *CELF1* and *KLC1* splice variants in the first human brain bank. The mRNA expression levels of brains of Japanese patients pathologically diagnosed with Alzheimer’s disease (AD) (n=10, shown in red) and those of control subjects (n=14, shown in blue) were measured by reverse transcription quantitative polymerase chain reaction (RT-qPCR) **a**. Expression levels of *CELF1* in AD and control brains. Long and short green bars indicate the mean and S.E. The mean expression level in the control group was normalized to 100. **b**. Expression levels of *CELF1* in brains with the risk allele C rs10838725 of *CELF1* (TC or CC: n=9) or no risk allele (TT: n=15). **c**. Correlation between *CELF1* levels and each splice variant of *KLC1* in the human brain. P values less than 0.01 (0.05/5 tests) according to Bonferroni correction for multiple testing were considered significant (continuous line).

### Alternative *KLC1* exons downstream of the CELF1 binding region were highly expressed in AD brains and those with low *CELF1* levels

To confirm and further elaborate the relationship between *CELF1* and *KLC1* in the human brain, we examined a large number of autopsied brains of different ethnicities using different methods. RNA-seq data from autopsied brains of American Caucasian AD (n=82), control (n=57), and progressive supranuclear palsy (PSP) groups (n=37) provided by AMP-AD Mayo^11^ were analyzed. Similar to the RT-qPCR analysis of Japanese autopsied brains (Fig. 2a), we observed lower *CELF1* levels in AD brains than those in control brains (Fig. 3a, p=0.023). In addition, the *CELF1* level was significantly lower (p=0.0002) in AD brains than in those of patients with PSP (n=37), which is a neurodegenerative disease with no Aβ pathology (Fig. 3b). This suggested that the low *CELF1* level observed in AD is not a common molecular mechanism in neurodegenerative diseases.

**Fig. 3.**
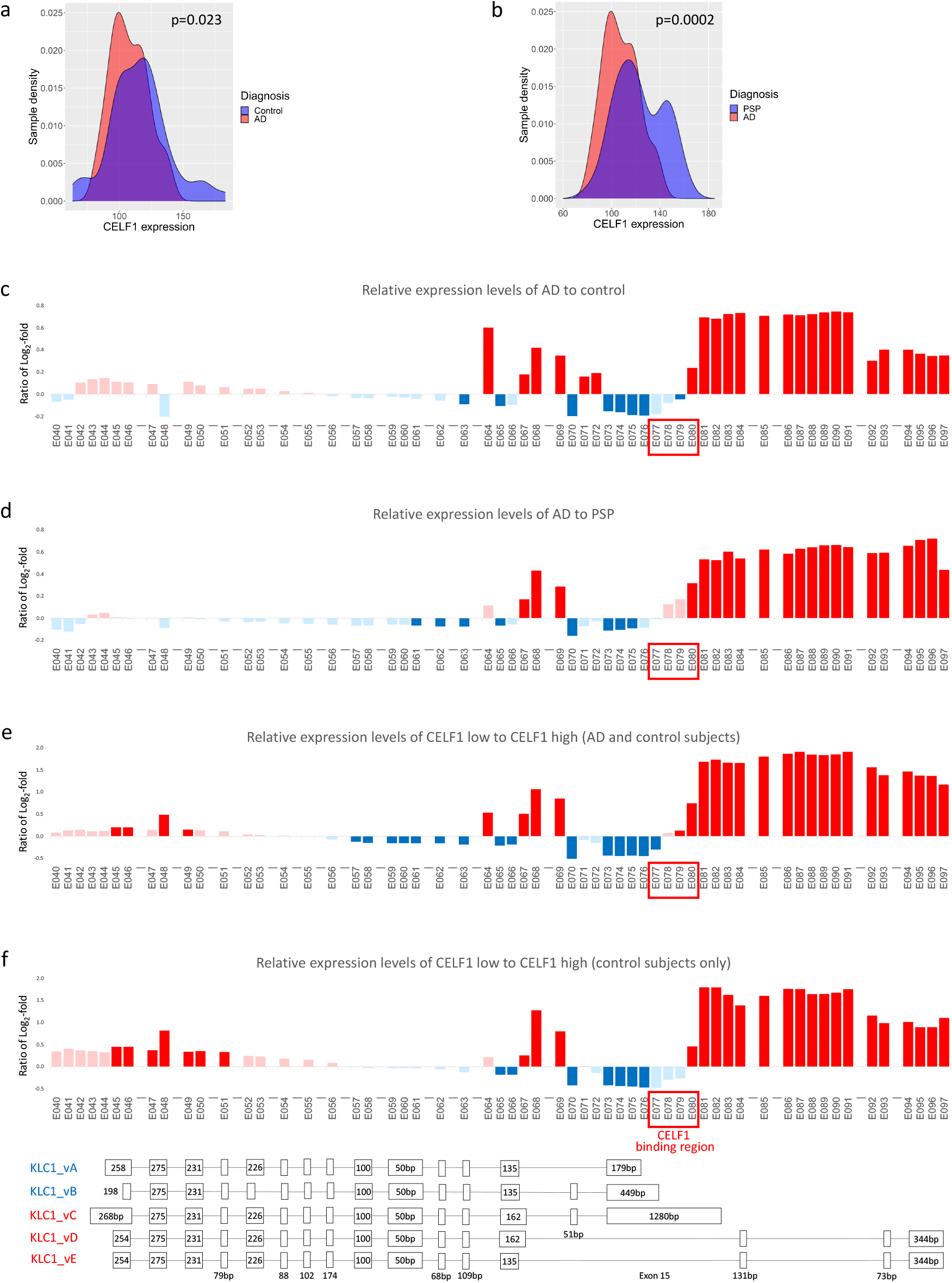
Expression levels of *CELF1* and *KLC1* exons in the second human brain. The American Caucasian RNA-seq dataset from AMP-AD Mayo was analyzed by 198 DEXSeq. **a**. Expression levels of *CELF1* (ENST00000395290) (X-axis) in Alzheimer’s disease (AD) (n=82, red) and control (n=57, blue) brains analyzed by DEXSeq. *CELF1* levels were significantly (p=0.023) lower in AD brains than those in control brains. **b**. The data are similar to those in a, but *CELF1* levels in samples from AD brains are compared with those from brains of neuropathologically confirmed progressive supranuclear palsy (PSP) patients lacking Aβ pathology (n=37, blue). *CELF1* levels were significantly (p=0.0002) lower in AD brains than those in PSP brains. **c–f** Relative expression levels of *KLC1* exons (feature IDs). The horizontal axis represents the *KLC1* exon (indicated by the feature ID used in DEXSeq). “-” indicates an intron spanning region. Red boxes indicate the CELF1 binding region (E077–080). The right end is the 3’ end. The vertical axis indicates the logarithm of the relative expression level (Log2). A dark color indicates an adjusted p<0.2; a light color indicates an adjusted p≥0.2. **c**. Expression levels of *KLC1* exons (feature IDs) in AD relative to control brains. Red and blue indicate high and low levels, respectively, in AD brains. **d**. Expression levels of *KLC1* exons (feature IDs) in AD brains relative to those from PSP patients. Red indicates high levels in AD brains; blue indicates high levels in PSP brains. **e**. Expression levels of *KLC1* exons (feature IDs) in the lower quartile relative to the higher quartile of *CELF1* in the combined AD and control samples. Red indicates high levels in the *CELF1* lower quantile; blue indicates low levels in the *CELF1* lower quantile. **f**. Expression levels of *KLC1* exons (feature IDs) in the lower quartile relative to the higher quartile of *CELF1* in control samples only. Below, the locations and sizes of the exons of the major KLC1 splice variants are shown with the variants positively (blue) and negatively (red) correlated with CELF1 levels in the human brain (see Fig. 2).

Because many *KLC1* exons have numerous alternative splice sites, we used DEXSeq^21^ to analyze RNA-seq data and calculated the expression levels of exons with multiple splice sites by dividing them into multiple feature IDs. The genomic location, expression levels, and statistics of each feature ID are shown in Supplementary Table 1. The expression levels of many exons (feature IDs) at the 3’ end^16^ (feature IDs E067–E097), where the major alternative exons are concentrated (Fig. 3 bottom), were significantly altered in AD brains compared with those in control brains (Fig. 3c). Feature IDs E084, E093, and E094–096, which are included in *KLC1_vE*, were elevated in AD samples, consistent with a previous report^10^, indicating that *KLC1_vE* measured by RT-qPCR was elevated in AD brains. The feature ID expression pattern in AD relative to PSP brains (Fig. 3d) was similar to that in AD relative to control brains (Fig. 3c), suggesting that this *KLC1* feature ID expression pattern is AD-specific rather than common to neurodegenerative diseases.

To investigate the relationship between this AD-specific KLC1 feature ID expression pattern and *CELF1* expression levels, we compared *KLC1* feature ID expression in brains with low and high *CELF1* expression levels (Fig. 3e). The expression of many *KLC1* feature IDs was significantly different in brains with low *CELF1* expression from that in brains with high *CELF1* expression, and the pattern was surprisingly similar to the AD-specific pattern. Furthermore, even when only control brains were analyzed, the *KLC1* feature ID expression pattern was still significantly altered in brains with low *CELF1* levels, and the expression pattern was similar to the AD-specific pattern (Fig. 3f). The control brain results only suggest that the AD-specific expression pattern of *KLC1* feature IDs in brains with low *CELF1* expression is not caused by an AD pathogenic mechanism independent of *CELF1* but is caused by a direct relationship between *CELF1* and *KLC1*. The low levels of feature IDs E073–076 and high levels of E080–097 caused by low *CELF1* levels (Fig. 3e and f) were consistent with the positive correlation of *KLC1_vA* and *vB*, levels with CELF1 levels and the inverse correlation of *KLC1_vC, vD* and *vE* levels with *CELF1* levels (Fig. 2c).

Notably, CLIP-seq showed that the CELF1 protein binds to feature IDs E077–E080 (red boxes in Fig. 3c–f), which are immediately upstream of the feature IDs that are elevated in both AD and low CELF1-expressing brains.

In summary, the DEXSeq analysis of RNA-seq data was consistent with the high *KLC1_vE* levels in AD brains^10^ and the correlation between major splice variants of *CELF1* and *KLC1* (Fig. 2c) shown by RT-qPCR. The DEXSeq results also showed that the expression pattern of *KLC1* exons (feature IDs) was highly similar in AD and low-*CELF1* brains and suggested that low *CELF1* levels upregulated the expression of *KLC1* exons downstream of the CELF1 binding site, resulting in high *KLC1_vE* levels.

### The control of *KLC1_vE* expression by *CELF1* was confirmed in cultured cells

Because CELF1 has been suggested to regulate *KLC1_vE* expression in the human brain, we performed cell culture experiments to directly confirm the causal relationship between the two molecules. We examined *KLC1* in three cell lines using three different methods: RT-qPCR (Fig. 4a and b), conventional RT-PCR (Fig. 4c and Supplementary Fig. 3), and exon arrays (Fig. 1a, b and Fig. 4d). In HEK293 cells, knockdown of *CELF1* significantly (37.5%–61.1%, 0.0016–<0.0001) increased *KLC1_vE* expression (Fig. 4a), and overexpression of *CELF1* significantly (27.4%, p=0.0008) decreased *KLC1_vE* expression (Fig. 4b). The relationship between CELF1 and *KLC1_vE* observed in HEK293 cells was consistent with that observed in the human brain, and the HEK293 cell experiment results confirm that *CELF1* expression levels determine *KLC1_vE* expression levels.

**Fig. 4.**
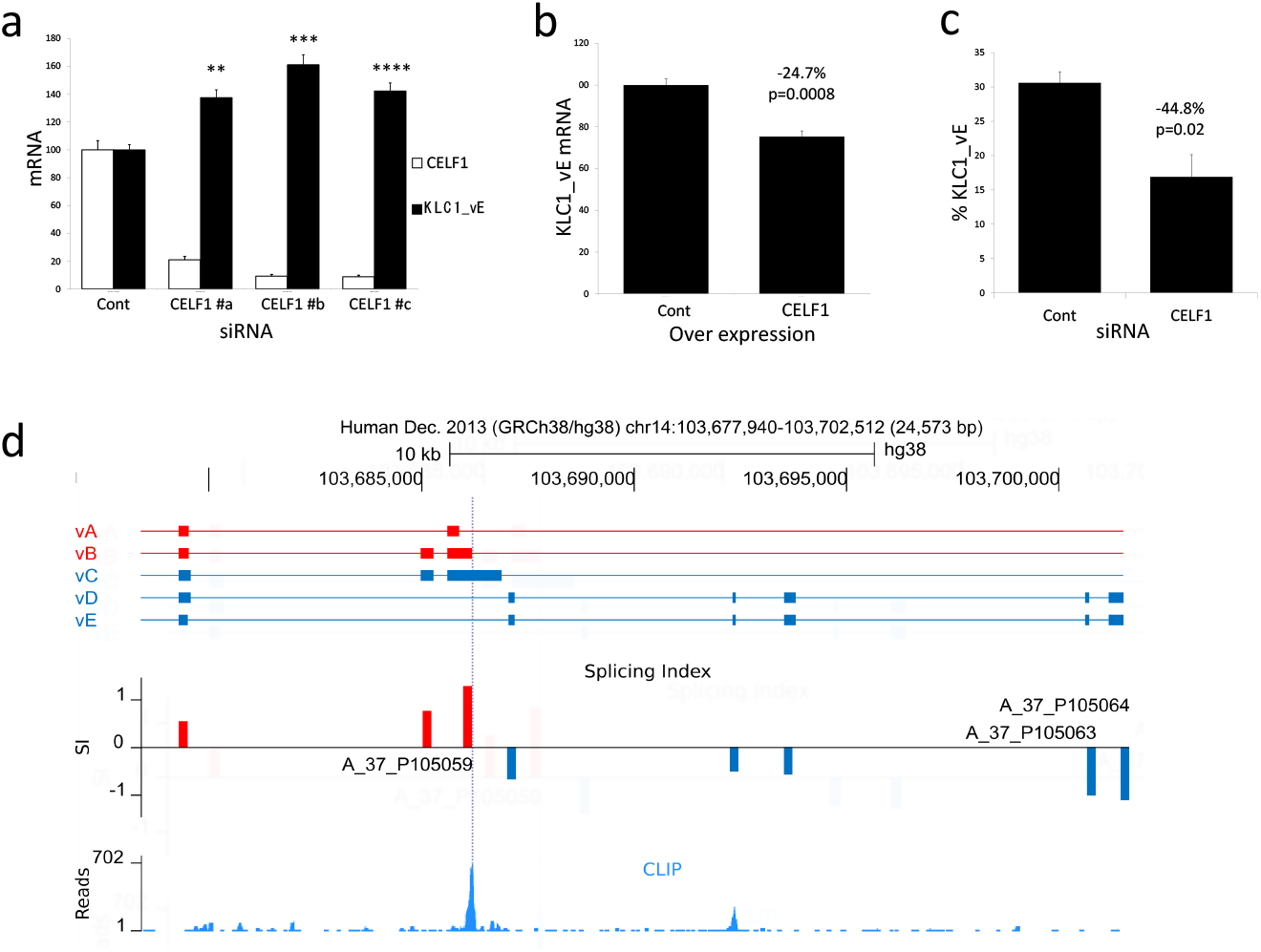
*CELF1* regulates *KLC1_vE* expression levels in HEK293 and HeLa cells. **a**. *KLC1_vE* mRNA levels in *CELF1* knockdown HEK293 cells measured by reverse transcription quantitative polymerase chain reaction (RT-qPCR). Suppression of *CELF1* by three siRNAs (n=4/treatment) induced *KLC1_vE* expression. ** p<0.01, *** p<0.001, **** p<0.0001 Dunnett’s multiple comparison test. Error bars indicate S.E. **b**. *KLC1_vE* mRNA levels in HEK293 cells overexpressing *CELF1* measured by RT-qPCR. Induction of *CELF1* (n=4/treatment) repressed *KLC1_vE* expression (−27.7%, p=0.0008). **c**. Percentage of *KLC1_vE* mRNA levels relative to all major variants of *KLC1* in HeLa cells measured by conventional RT-PCR. Repression of *CELF1* decreased *KLC1_vE* expression (−45.1%, p=0.02, three independent experiments). **d**. Changes in the expression of alternative exons of *KLC1* by *CELF1* knockdown in HeLa cells shown by exon array analysis. Upper, alternative exons on the 3’ side of the major splice variants of *KLC1* are shown. Note that because of the alternatively spliced donor sites, the size of the left exon differs between *vC* and *vD* and those of the other variants. Color code, red and blue bars indicate variants that are positively and negatively correlated, respectively, with *CELF1* levels in the human brain (see Fig. 2). Middle, changes in expression of the alternative exons of *KLC1* by *CELF1* knockdown are shown. The vertical axis shows the splicing index (SI) of exon array probes (*CELF1* knockdown samples compared with control samples). The indicated probe IDs are three of the 10 probes most substantially altered by *CELF1* knockdown shown in Fig. 1a. Red and blue indicate higher and lower inclusion levels following *CELF1* knockdown. Bottom, the CELF1 binding region identified by cross-linking immunoprecipitation sequencing is shown. The major CELF1 cross-linking immunoprecipitation sequencing peak (dotted purple line) delineates the boundary between increased or decreased *KLC1* exon expression.

We also examined *KLC1_vE* regulation in SH-SY5Y (Supplementary Fig. 3) and HeLa cells (Fig. 4c–d) and found that, in contrast to HEK293 cells and the human brain, *KLC1_vE* was significantly reduced in SH-SY5Y (p=0.0005) (Supplementary Fig. 3) and HeLa cells (p=0.02) (Fig. 4c) by *CELF1* knockdown. Therefore, we performed a detailed reanalysis of the exon array of *CELF1* knockdown HeLa cells shown in Fig. 1a. Fig. 4d shows the genomic locations and expression levels of *KLC1* probes, including the three probes included in the top 10 *CELF1* knockdown expression changes (Fig. 1a), and the CLIP-seq results of the CELF1 protein (Fig. 1b). *CELF1* knockdown increased *KLC1* exon expression in the CELF1 binding region and that of exons immediately upstream of this region and decreased expression of all exons immediately downstream of the CELF1 binding region. Although the direction of CELF1 regulation was opposite to that of the human brain, the switch in the increase and decrease of *KLC1* exons upstream and downstream of the CELF1 binding region was consistent with that in the human brain.

Taken together, these results suggest that *KLC1_vE* expression is not solely determined by CELF1, but that cell- and tissue-specific factors determine the direction of regulation. More importantly, CELF1 regulation of *KLC1_vE* expression was observed in all cells and tissues examined.

### *Klc1_vE* expression levels determine the propensity for Aβ pathology in each mouse strain

Current and previous studies^10^ have uncovered novel molecular pathways in AD that involve the risk gene *CELF1* in the following pathway: *CELF1*→*KLC1_vE*→Aβ pathology. To investigate whether the entire molecular pathway of *CELF*→*KLC1_vE*→Aβ pathology can occur within the same cell, we examined a single-cell RNA sequencing database. PanglaoDB^20^ (https://panglaodb.se/index.html) showed that the Aβ precursor protein (*APP), CELF1*, and *KLC1* are co-expressed mainly in neurons (Supplementary Fig. 1b).

In addition to identifying the first half of the pathway (*CELF1*→*KLC1_vE*), we also performed additional experiments to reconfirm the previously elucidated second half of the pathway (*KLC1_vE*→Aβ). Our unique use of transcriptomics focusing on mouse genetic backgrounds, instead of comparing *APP* transgenic and non-transgenic mice, revealed higher *Klc1_vE* levels in C57BL/6 and SJL mice than those in DBA/2 mice because of the differences in the *Klc1* allele, and the increased *Klc1_vE* level accelerates Aβ pathology in these two mouse strains^10^.

In addition to our group, six groups have reported differences in Aβ pathology in various mouse strains^22,23,24,25,26,27,28^. Except for our study, these studies have not identified genes that modulate Aβ pathology. Therefore, we examined whether the mouse strain effects on Aβ found in these studies can be explained by *Klc1*. Although various AD model mice (Tg2576^29^, APP23^30^, R1.40^31^, TgCRND8^32^, APB (JAX MMRRC Stock# 034832), and APPPS1^33^) were used, the order of mouse strain effects (from high to low) on Aβ pathology was consistent in the six research groups (Fig. 5a), except for a single study^28^. Thus, we analyzed seven mouse strains (C57BL/6, SJL, 129S, A/J, DBA/2, C3H, and FVB) used in these consistent studies. We compared *Klc1* SNPs in these seven mouse strains using the Mouse Phenome Database^34^ (RRID:SCR_003212, https://phenome.jax.org/). The available *Klc1* SNPs for all seven strains are shown in Fig. 5b, and the full SNP set with RefSNP (rs) numbers is shown in Supplementary Table 2. Although these seven mouse strains belong to various phylogenetic groups^35^ (Supplementary Table 3), these SNPs revealed that high Aβ mouse strains (C57BL/6, SJL, and 129S1) share a similar *Klc1* allele, while low Aβ mouse strains (A/J, DBA/2, C3H, and FVB) share another *Klc1* allele, as previously shown in part^10^. To reconfirm that the *Klc1_vE* expression level affects Aβ pathology in the mouse brain^10^, we measured the *Klc1_vE* mRNA level in these seven mouse strains. We observed higher *Klc1_vE* expression levels in high Aβ mouse strains at 3 months (Fig. 5c) and 12 months of age (Supplementary Fig. 4) than those in low Aβ mouse strains, as expected. To determine whether *Klc1_vE* was decreased in DBA/2 mice throughout the lifespan, we examined brains from mice aged 4 to 64 weeks and observed that *Klc1_vE* levels were lower in DBA/2 mice than those in C57BL/6 mice at all ages (Supplementary Fig. 5). These results reconfirm that the severity of Aβ pathology in mice is determined by the *Klc1_vE* expression level.

**Fig. 5.**
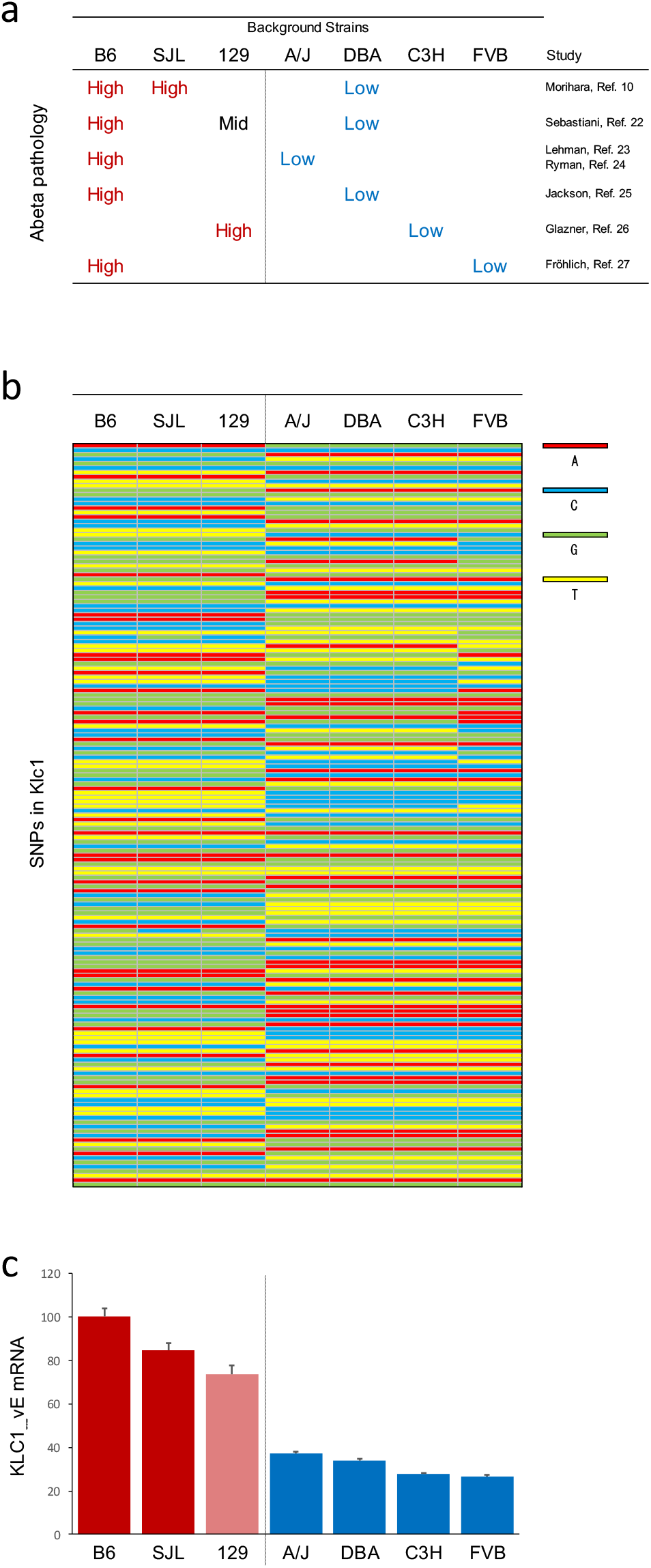
Mouse strains with enhanced amyloid-β (Aβ pathology share a common *Klc1* allele and express higher *Klc1_vE* levels, while those with suppressed Aβ pathology share a different *Klc1* allele and express lower *Klc1_vE* levels. **a**. The severity of Aβ pathology in each mouse strain as reported by six groups. In each study, mouse strains with more severe Aβ pathology are shown as “High” in red, those with milder pathology are shown as “Low” in blue, and moderate pathology is indicated as “Mid” in black. **b**. *Klc1* single nucleotide polymorphisms (SNPs) in seven mouse strains. Color bars indicate 168 SNPs in the *Klc1* region (chromosome 12: 111,768,636–111,803,435 bp; build 38) of the seven mouse strains. Mouse strains with high Aβ levels, including C57BL/6, SJL, and 129S1, share a similar *Klc1* allele, while those with low Aβ levels, including A/J, DBA/2, C3H, and FVB, share another *Klc1* allele. **c**. Brain expression levels of *KLC1_vE* mRNA in the seven mouse strains at 3 months of age. *Klc1* mRNA levels in each high Aβ strain (B6, SJL, and 129, shown by red and pink bars) were significantly higher (Tukey-Kramer HSD test, p<0.0001) than those in low Aβ strains (A/J, DBA, C3H, and FVB, shown by blue bars). The expression level of B6 mice was set to 100. Error bars indicate S.E. (n=6–8 for each strain. The total number of animals was 54.)

## Discussion

This study revealed that CELF1, which is encoded by a gene located at a risk locus for AD^8^, regulates expression of the Aβ pathology-driving gene product *KLC1_vE*. The connection between *CELF1* and *KLC1* was revealed by integrating two functional omics approaches, the marked change in *KLC1* splicing in *CELF1* knockdown cells shown by an exon array (Fig. 1a) and binding of CELF1 protein to *KLC1* RNA shown by CLIP-seq (Fig. 1b), which led to subsequent studies (Fig. 6). Analysis of two brain banks, each with a different ethnicity, using different methods revealed strong regulation of *KLC1* splicing by CELF1 (Fig. 2 and 3). The *CELF1* level was low in AD brains, and alternative *KLC1* exons on the 3’ side of the gene exhibited high levels, which resulted in high levels of *KLC1_vE*. These changes were not observed in brains of patients with PSP, another neurodegenerative disease, and may be AD-specific. In addition, the AD-specific expression pattern of alternative *KLC1* exons was similar to that observed in brains with low *CELF1* levels, indicating that low *CELF1* levels cause AD-specific *KLC1* splicing, resulting in high *KLC1_vE* levels. Strikingly, CLIP-seq showed that the CELF1 binding site on *KLC1* RNA is immediately upstream of the exons that are elevated in both AD and low CELF1-expressing brains (Fig. 3c–f). Although the direction of the regulatory effect depended on the cell type, CELF1 regulated *KLC1* splicing, including *KLC1_vE*, in all three types of cultured cells examined (Fig. 4). Previously reported regulation of Aβ pathology by *KLC1_vE* was reconfirmed in our analysis of seven mouse strains, in which the strains prone to Aβ pathology had high *Klc1_vE* expression levels and shared a common *Klc1* allele (Fig. 5). Our findings indicate that low expression of the risk gene *CELF1* increases the inclusion of *KLC1* exons downstream of the CELF1 binding region, resulting in increased *KLC1_vE* expression and consequently driving Aβ pathology as a novel molecular pathway in the development of AD.

**Fig. 6.**
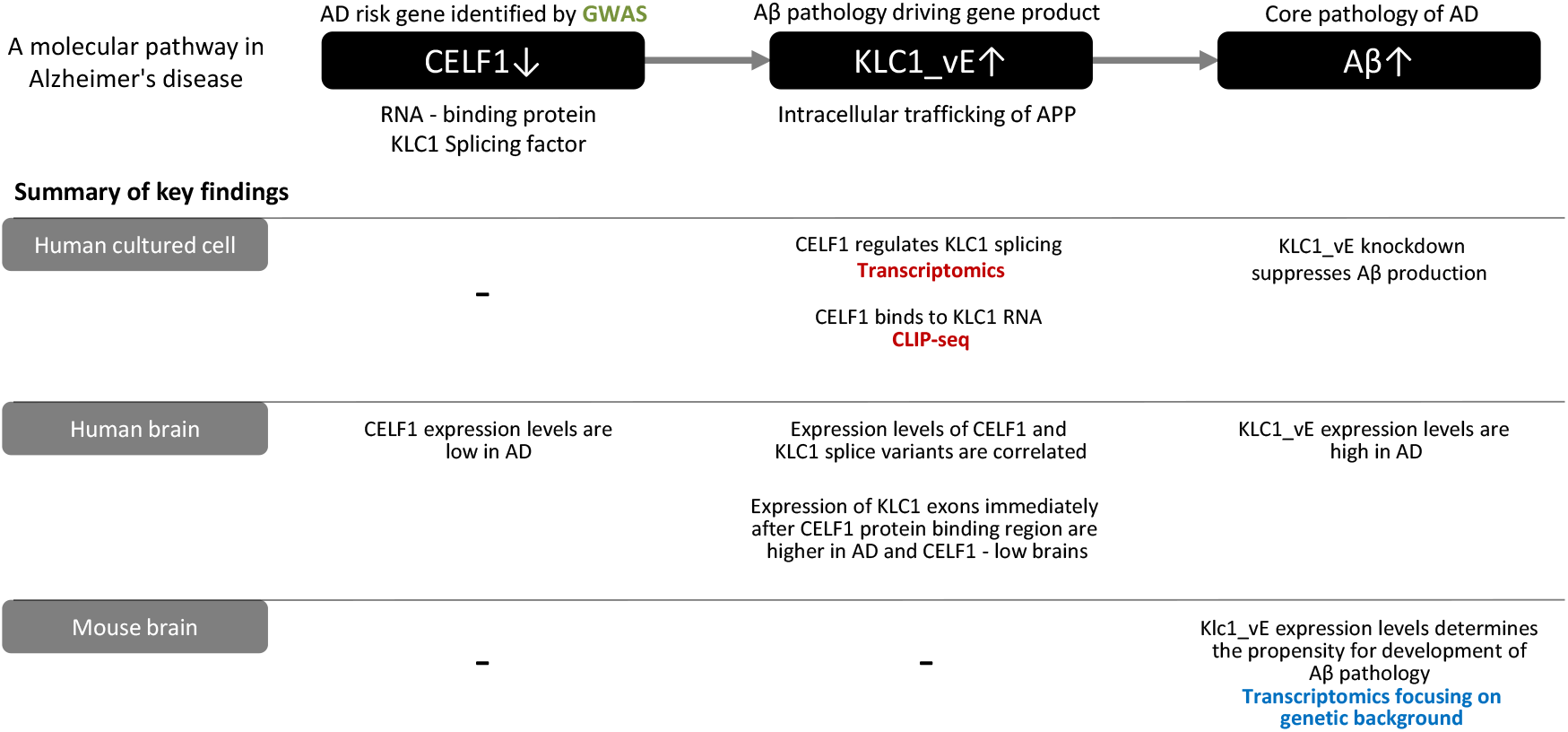
Hypothesis-free omics approaches identified a molecular pathway for Alzheimer’s disease (AD) development, which was confirmed in cultured cells, autopsied human brains, and mouse experiments. The omics data that identified relationships are shown in color. Green: The *CELF1* locus was associated with AD in genome-wide association studies (Ref. 3–6). Red: Transcriptomic studies using *CELF1* knockdown revealed strong regulation of KLC1 by CELF1; cross-linking and immunoprecipitation sequencing identified *KLC1* RNA as a binding partner of the CELF1 protein and revealed its binding site. Blue: Transcriptomic studies focusing on mouse background genes revealed that *Klc1_vE* expression and the *Klc1* allele are responsible for the differences in Aβ accumulation among mouse strains (Ref. 10).

Including our previous studies^10^ and those of other groups^8^, a total of four different types of omics led to the discovery of the *CELF1*→*KLC1_vE*→Aβ pathway (Fig. 6). Two unexpected connections, regulation of *KLC1* splicing by CELF1 and the role of *KLC1_vE* in Aβ pathology^10^, were discovered by functional omics, such as transcriptomics and CLIP-seq. The advantage of these functional omics methods over GWASs is that they reveal the identity and interactions of specific genes, suggesting a possible functional relationship. However, functional analysis, especially using mRNA expression levels, has some potential caveats. Regulation of expression and splicing is complex, is modulated by many molecules, and can be tissue- and cell-type-specific^36,37^. In our experiments, the control exerted by CELF1 over *KLC1* splicing led to opposite effects on expression levels of *KLC1* splice variants in human brains and two of three types of cultured cells. As neuronal splicing is especially complex^38^, factors other than CELF1 may also regulate *KLC1* splicing, and further studies are needed to fully understand the *KLC1* splicing mechanism. In a similar example, the relationship between *CELF1* expression and cancer patient survival is variable in different cancer types^14^. When conducting functional genomics for disease research, it is important to confirm the final conclusion of the functional expression analysis at the site of the disease lesion, which in the case of AD is the brain.

KLC1 is part of the Aβ production mechanism^10,19,39,40^ and is responsible for intracellular trafficking, including axonal transport^19,41,40^. A recent analysis of Alzheimer’s Disease Neuroimaging Initiative data reported that an SNP of *CELF1* may cause abnormalities in axonal tracts in the hippocampus^42^. *CELF1, KLC1*, and *APP*, which undergoes proteolysis to form Aβ, coexisted in the same cells, which mainly included neurons (Supplementary Fig. 1b). KLC1 is an adapter protein that forms a complex with the motor protein kinesin heavy chain and transports various cargoes to subcellular locations^19^. APP is carried by KLC1/Kinesin-I^43,44,40^, and knockdown of total KLC1^39,10^ or *KLC1_vE* ^10^ suppresses Aβ production. However, Aβ pathology is exacerbated in heterozygous *Klc1* knockout mice^41^. Thus, the expression of *Klc1_vE* in heterozygous *Klc1* knockout mice may explain the worsening of Aβ pathology.

The complex alternative splicing of the 3’ exon of *KLC1* results in a variety of splice variants. Complex splice variants and the exon structure are both highly conserved among species^16^, suggesting that splicing of the 3’ exon is functionally important. We found that CELF1 regulates the expression of these important exons. It is speculated that splicing at the C-terminus of the KLC1 protein regulates the selection of cargo^45^ and its transport destination^46^. For example, calsyntenin-1 (alcadein-α) is involved in APP transport by binding to KLC1 and APP and consequently affects Aβ production^40^. KLC1_vE, unlike other splice variants, has been reported to be unable to bind Calsyntenin-1^47^. Further studies are expected to elucidate the distinct function of KLC1_vE.

This study not only revealed the function of CELF1 in AD but also confirmed the importance of KLC1_vE in AD. When a master regulator gene, which trans-regulates the expression of many genes, is a disease-associated gene, the effect size on the disease is usually small. CELF1 is an RNA-binding protein that regulates the expression of many genes. Many of the master regulator genes are “peripheral genes” that are not directly involved in common complex diseases but regulate “core genes” that are directly involved in disease development^48,49^. CELF1 regulates the expression of many genes as an RNA-binding protein and may be a “peripheral gene” that also trans-regulates the splicing of *KLC1*, a “core gene” in AD. Understanding the role of the “core gene” *KLC1* may lead to the development of new therapies for AD.

It is noteworthy that a decrease in Aβ pathology was achieved not by genetic manipulation but by naturally occurring low levels of *Klc1_vE* that are shared by many mouse strains. Many reports have described the suppression of AD pathology by genetic manipulation in mice; however, the therapeutic target genes often have unknown physiological effects in addition to their effects on AD. In the case of genetically engineered mice, the side effects caused by inhibition of these unknown physiological effects are often overlooked. In fact, so-called mechanism-based side effects, which are inevitable when this therapeutic target is used, may go unnoticed until large-scale clinical trials are conducted^50,51^. It is highly likely that *KLC1_vE* also transports other cargo that is yet to be identified. However, marked reductions in Aβ pathology have been achieved via naturally occurring low *Klc1_vE* levels in commonly used mouse strains. This indicates that the risk of mechanism-based side effects when *KLC1_vE* is used as a therapeutic target is minimal.

In summary, we found that CELF1, an RNA-binding protein, binds to *KLC1* RNA. We also demonstrated that low CELF1 levels increase the expression of *KLC1* exons immediately downstream of the CELF1 binding site, resulting in increased *KLC1_vE* mRNA levels. These findings revealed a new pathway for the pathogenesis of AD, in which low levels of CELF1, located in a risk locus for the disease, increase *KLC1_vE* expression and promote Aβ pathology. Importantly, this pathway is supported by human and mouse genetic data and was derived from various types of hypothesis-free omics approaches. This study may be a useful example to elucidate the function of risk genes for many complex diseases.

## Data Availability

The CELF1 CLIP-seq data have been published (Genom Data. 2016 Apr 19;8:97-103. doi:10.1016/j.gdata.2016.04.009).
The exon array experiments are available as a preprint (https://doi.org/10.1101/373704), and the raw microarray data are available in GEO as data set GSE11898

## Acknowledgment

This work was supported by the Strategic Research Program for Brain Sciences (to T.M.), KAKENHI (C) (23591706 and 26461747 to T.M.), Takeda Science Foundation (to T.M.), SENSHIN Medical Research Foundation (to T.M.), Restar Communications Corporation (to M.I. and T.M.), the departmental committees of the Ligue Contre le Cancer du Grand Ouest 22, 29, 35 (to L.P.), the Japanese Brain Bank Network for Neuroscience Research (to H.A.), and the Natural Sciences and Engineering Research Council of Canada (2018-03945 to M.A.S.).

## Materials and methods

### Omics analyses in HeLa cells

We have previously described the results of exon array experiments using Agilent (Santa Clara, California, USA) SurePrint G3 human exon array probes. Hybridized RNA was extracted from HeLa cells previously treated with siRNAs to repress *CELF1, ELAVL*, or both proteins or a control siRNA^14^. We examined the control siRNA and *CELF1* siRNA datasets to identify the most highly differentially expressed exons. The CLIP-seq results have been previously published^15^. We used the UCSC genome browser to reveal CELF1 binding to *KLC1* RNA.

### Human brains

The same hippocampi that were previously analyzed^10,52^ were reanalyzed. Pathologically diagnosed autopsied brains were obtained from the brain bank of the Choju Medical Institution of Fukushimura Hospital. The RNA Integrity Numbers (RINs) of 10 AD and 14 control mRNAs were all greater than 7. The protocol was independently approved by the local ethics committees of Osaka University and Fukushimura Hospital.

### Genotyping of CELF1

An AD risk SNP (rs10838725) in the *CELF1* region was genotyped using a custom TaqMan(R) SNP Assay (Assay ID: AHWSKQD, Thermo Fisher Scientific, Waltham, Massachusetts, USA). Forward primer sequence: TGGAGACTGAGGCACGAGAA, Reverse primer sequence: TGGGTTATAGATAGCATTGGGAAAC, VIC/FAM sequence: AGCCAACAT/CTGCACCAC

### Quantitative PCR

Splice form-specific qPCR assays were designed using Primer Express (Thermo Fisher Scientific), as described previously^10^. The forward primer and TaqMan MGB probe sequences for human *KLC1_vA, vB, vD*, and *vE* were TCTCGTAAACAGGGTCTTGACAATG and ATGACCCTGAGAACAT, respectively.

The reverse primer sequences were *KLC1_vA*: ACGGGCGGCTAGGCTTC; *KLC1_vB*: CGAGCTTCATTTTCCTCATTTCC; *KLC1_vD*: GCCATCCCCGTTCCACTC; and *KLC1_vE*: TTTAAAGATCCAGTGCCATCTTCC. For human *KLC1_vC*, the forward primer sequence was ACCAGCCCGAGCTTCATTT, the reverse primer sequence was ACGGAGGGGAGGAAGTGAGT, and the TaqMan MGB probe sequence was TCCTCATCCCGTTCCA. The total levels of human *KLC1* were measured using a TaqMan(R) Gene Expression Assay (Hs00194316_m1, Thermo Fisher Scientific) via detection of the exon 3 and 4 boundary, where exon usage was stable according to the DEXSeq results. The level of each splice variant was normalized using the total *KLC1* level. Human *CELF1* mRNA levels were measured using a TaqMan(R) Gene Expression Assay (Hs00198069_m1, Thermo Fisher Scientific).

For mouse *Klc1_vE*, the forward primer sequence was GGGTCTTGACAATGTTCACAAAC, the reverse primer sequence was GATCCAGTGCCATCTTCCTCC, and the TaqMan MGB probe sequence was TCCGTCAGGGCCACT.

The TaqMan(R) Gene Expression assay (Mm00492936_m1) with primers for exons 3 and 4 was used to detect the total level of mouse *Klc1*. The expression level of *Klc1_vE* was normalized to that of total *Klc1*.

### Conventional RT-PCR

Conventional RT-PCR was performed using RNAs from cells previously depleted of *CELF1* by siRNA treatment. Reverse transcription was performed with random primers and Superscript II RT (Thermo Fisher Scientific). PCR was performed with the following primers: Forward: AACAGAGGGTGGCAGAAGTG; Reverse R1: GCAGCACTGGAGAGAGAAGG; Reverse R2: TCCCTTCAGCTTCCTAACCA. Relative amounts of *KLC1-vE* were determined by calculating the intensity of the vE PCR product divided by the sum of the intensities of the five PCR products.

### Single-cell RNA sequencing database

Co-expression of *CELF1, KLC1*, and *APP* was analyzed in the PanglaoDB^20^ (https://panglaodb.se/index.html) on April 12, 2021 under the following conditions: species: mouse and human, tumor/cancer samples, cell lines were not included, non-adult and non-primary samples were not included.

### Analysis of the human temporal cortex RNA-seq data from the AD Knowledge Portal

We analyzed the RNA-seq data using the DEXSeq package to detect differential exon usage^53^. The RNA-seq data were obtained from the AD Knowledge Portal (syn3163039) as BAM files, and the raw sequencing data were mapped to the reference sequence (GRCh38.77) using SNAPR software (arXiv technical report 1111.5572v1, Nov. 2011). These RNA-seq data were obtained from the temporal cortex of subjects (AD patients (n=84), PSP patients (n=84), and controls (n=80)) from the Mayo Clinic Brain Bank and Banner Sun Health Research Institute. Among the subjects, 82 AD patients, 84 PSP patients, and 57 controls with an RIN of 7 or greater were included. We included only PSP patients with Thal amyloid phase 0 (n=37) to exclude patients with AD pathology. Detailed processing and sample quality control are described by Allen *et al* ^54^.

We counted the number of reads that overlapped genes in the reference transcriptome annotations (GRCh38.77) using featureCounts^55^. To quantify exon expression, we first created an exon annotation file (GFF) using the reference transcriptome annotations and dexseq_prepare_annotation2.py script from the DEXSeq package. We then used the aligned RNA-seq BAM files from the gene expression quantification and featureCounts to count the number of reads overlapping each exon. We estimated relative exon usage fold changes between the corresponding groups using an estimate function (ExonFoldChanges) of the DEXSeq package and output the results using the DEXSeqResults function. The DEXSeq package accounts for the case in which the exon boundaries of multiple transcript variants of a gene do not match and divides the exon into two or more parts. These are called “counting bins” and are identified by the “feature IDs”.

We obtained the expression levels of *CELF1* (ENST00000395290) from MayoRNAseq_RNAseq_TCX_covariates.csv in the AD Knowledge Portal. SNAPR software, which is an RNA sequence aligner, was used to align sequencing reads from Illumina HiSeq 4000 sequencers to the GRCh38 reference and Ensembl v77 gene models and generate read count data. The read count data were subsequently normalized using edgeR to calculate the counts per million mapped reads.

### Cultured cell experiments

We performed three independent *CELF1* knockdown experiments. Using the BLOCK-iT RNAi Designer (Invitrogen), we developed three *CELF1* Stealth siRNAs. The siRNA sequences and amounts were as follows:

*CELF1* #a siRNA: CACAGACGGCUAUCAAGGCAAUGCA (10 pmol/well)

*CELF1* #b siRNA: AGAAGAUUCUGGUUGUACAGAGUGG (3 pmol/well)

*CELF1* #c siRNA: CCACCCAGACCAACCAGAUCUUGAU (10 pmol/well)

One day before transfection, HEK293 cells were plated at 1.0×10^5^ cells/six-well culture plate. *CELF1* siRNAs were mixed with RNAiMAX (Invitrogen) and Opti-MEM Reduced Serum Medium (Gibco). RNA was isolated 72 h after transfection (RNeasy Mini Kit, Qiagen).

We also performed three independent *CELF1* overexpression experiments. One day before transfection, 5.0×10^5^ HEK293 cells were plated onto a six-well plate. HEK293 cells were transfected with 1.75 µg of human *CELF1* (RefSeq NM_001025596) or an empty pTarget plasmid (Promega) using 2.5 µl of lipofectamine LTX and Plus reagent (Invitrogen) according to the manufacturer’s protocol (n=4 per group). RNA was isolated 24 h after transfection (RNeasy Mini Kit, Qiagen).

### *Klc1* SNPs in seven mouse strains

The SNPs in the *Klc1* region (chromosome 12: 111,758,962–111,807,808, GRCm38 mm10) were obtained from the Mouse Phenome Database^34^ (https://phenome.jax.org/) for SJL mice on August 22, 2011, and for C57BL/6J, 129S1/SvImJ, A/J, DBA/2J, C3H/HeJ, and FVB/NJ mice on February 2, 2020. A total of 1703 *KLC1* SNPs that had been shown to vary in one or more of the seven mouse strains are shown in Supplementary Table 2 along with the RefSNP (rs) numbers. Of these, 168 SNPs that were available in all seven strains are shown in Fig. 5b.

### Mouse brain samples

Mouse brains were dissected, and RNA was extracted as previously described^10^. To minimize variability caused by aging, dissections were performed within 3 days of the scheduled time point for dissection. Before dissection, mice were perfused with 15–20 mL of 0.05 M tris-buffered saline (pH 7.2–7.4) containing a Protease Inhibitor Mixture (P2714; Sigma). Dissected brains were snap-frozen in liquid nitrogen, and the hippocampus was analyzed. All animal procedures were performed according to the protocols approved by the Osaka University Animal Care and Use Committee.

### Statistical analysis

Statistical analysis other than omics studies was performed using JMP Pro 14.3.0 (SAS Institute Japan, Tokyo, Japan). Data are presented as the mean ± s.e.m., and two-sided p values <0.05 were considered statistically significant, unless otherwise specified.

