## Supplemental Table 3 and Fig 1-5 for "An Alzheimer’s disease pathway uncovered by functional omics: the risk gene *CELF1* regulates *KLC1* splice variant E expression, which drives Aβ pathology"

**Supplementary Table 3** and **Supplementary Fig. 1-5**

An Alzheimer's disease pathway uncovered by functional omics:

the risk gene *CELF1* regulates *KLC1* splice variant E expression,

which drives Aβ pathology

Masataka Kikuchi^1,*^, Justine Viet^2,*^, Kenichi Nagata^3^, Masahiro Sato^4^, Géraldine David^2^, Yann Audic^2^, Michael A. Silverman^5^, Mitsuko Yamamoto^4^, Hiroyasu Akatsu^6,7^, Yoshio Hashizume^8^, Kyoko Chiba^9^, Shuko Takeda^10,11^, Shoshin Akamine^12,13^, Tesshin Miyamoto^4^, Ryota Uozumi^4^, Shiho Gotoh^4^, Kohji Mori^4^, Manabu Ikeda^4^, Luc Paillard^2,#^, Takashi Morihara^4,14,#^

^1^Department of Genome Informatics, Graduate School of Medicine, Osaka University, Suita, Japan.

^2^Univ Rennes, CNRS, IGDR (Institut de Génétique et Développement de Rennes), UMR 6290, F-35000 Rennes, France.

^3^Department of Functional Anatomy and Neuroscience, Graduate School of Medicine, Nagoya University, Nagoya, Japan.

^4^Department of Psychiatry, Graduate School of Medicine, Osaka University, Suita, Japan.

^5^Department of Biological Sciences, Centre for Cell Biology, Development, and Disease, Simon Fraser University, Burnaby, Canada.

^6^Department of Community-based Medical Education, Graduate School of Medicine, Nagoya City University, Nagoya, Japan.

^7^Choju Medical/Neuropathological Institute, Fukushimura Hospital, Toyohashi, Japan.

^8^Institute of Neuropathology, Fukushimura Hospital, Toyohashi, Japan.

^9^Frontier Research Institute for Interdisciplinary Sciences (FRIS), Tohoku University, Sendai, Japan.

^10^Department of Clinical Gene Therapy, Graduate School of Medicine, Osaka University, Suita, Japan.

^11^Osaka Psychiatric Medical Center, Osaka Psychiatric Research Center, Hirakata, Japan.

^12^Department of Mental Health Promotion, Graduate School of Medicine, Osaka University, Suita, Japan.

^13^Health and Counseling Center, Osaka University, Toyonaka, Japan.

^14^Center for Twin Research, Graduate School of Medicine, Osaka University, Suita, Japan.

*co-first authors

^#^co-last authors

(**Supplementary Table 1 and 2** are in the separate Excel files.)

**Supplementary Table 3** Mouse phylogenic tree groups

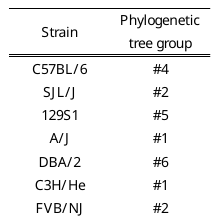

This table was based on “An efficient SNP system for mouse genome scanning and elucidating strain relationships.” Genome Research 2004 v14 p1806, where 102 mouse strains were organized into seven groups.

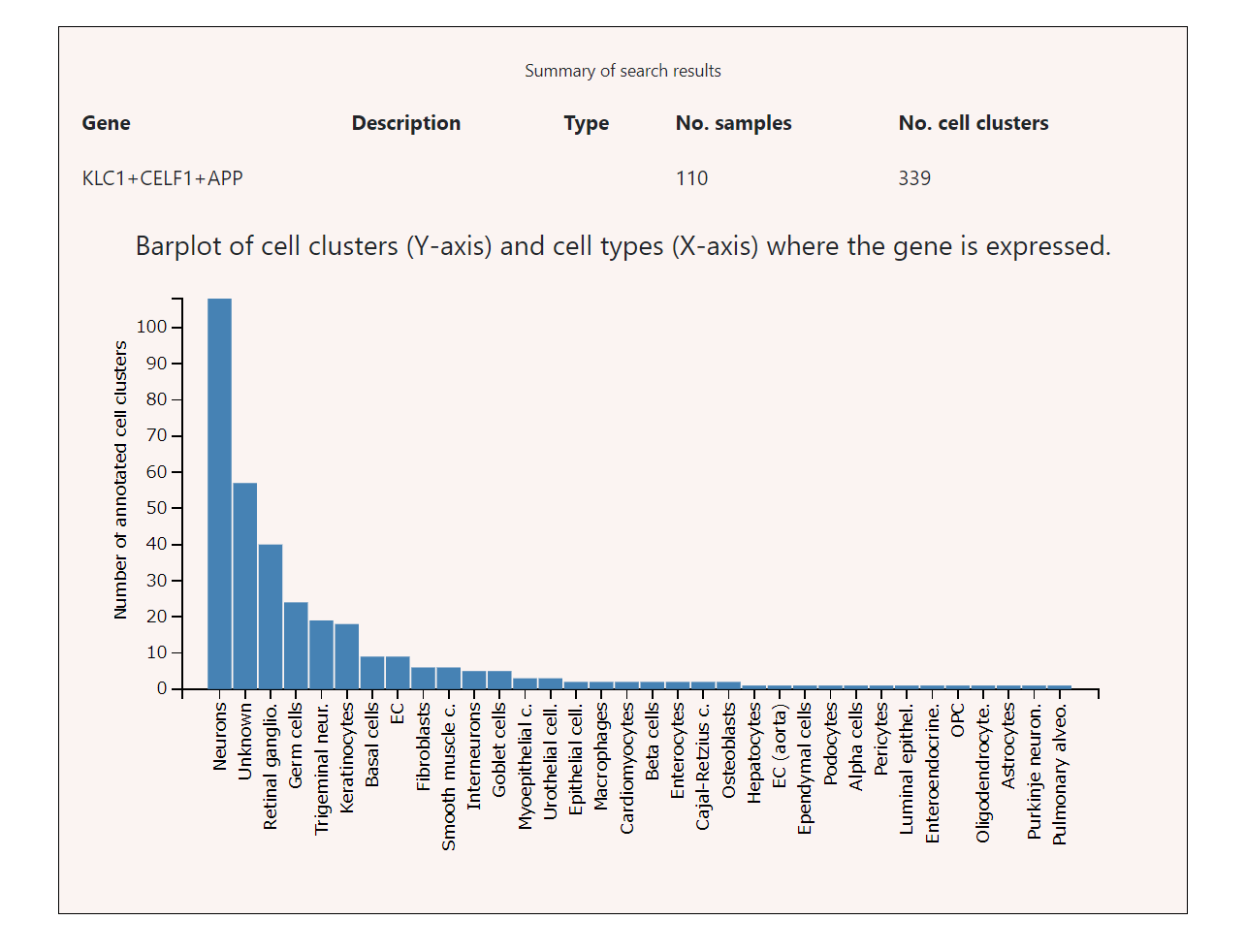
**a.**

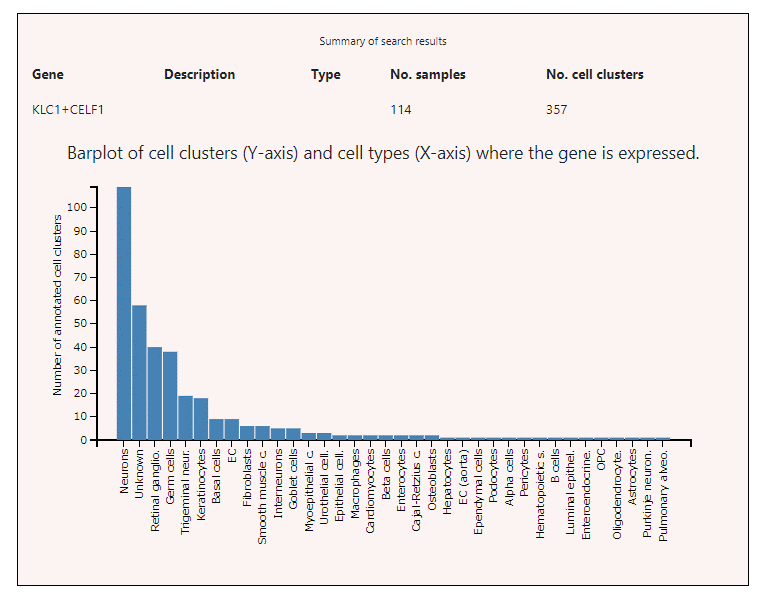

b.

**Supplementary Fig. 1 The number of cell clusters in which *CELF1, KLC1,* and *APP* were co-expressed for each cell type**

The single-cell RNA sequencing database PanglaoDB (<https://panglaodb.se/>) categorizes cell clusters on the basis of the single-cell gene expression profile. PanglaoDB is a database containing information from more than 1054 single-cell experiments using more than 4 million cells from a wide range of tissues and organs in humans and mice. After each cell was clustered on the basis of gene expression, each cell cluster was annotated using the marker gene sets of a cell type. Only clusters with at least 10 cells were used in the analyses. PanglaoDB provides the number of cell clusters where genes are expressed.

**a.** The cell clusters where *CELF1* and *KLC1* were co-expressed were primarily classified as neurons.

**b.** The cell clusters where *CELF1, KLC1,* and *APP* were co-expressed were primarily classified as neurons.

**Supplementary Fig. 2 Correlation between mRNA levels of *CELF1* and *KLC1* splice variants in control human brains**

The solid lines indicate a significant correlation. Dashed lines indicate no significant correlation. P values less than 0.01 (0.05/5 tests) according to Bonferroni correction for multiple testing were considered significant. n=14. The data are the same as those in Fig. 2c, but only control human brains are included.

**Supplementary Fig. 3 *KLC1_vE* expression quantified by conventional reverse transcription-polymerase chain reaction (RT-PCR) assays in SY5Y cells with *CELF1* knockdown**
**a.** Genomic structure of the *KLC1* gene and its five main variants. The positions of the PCR primers are indicated (one forward primer, F, and two reverse primers, R1 and R2). The size of the exon targeted by the forward primer differs between *vC* and *vD* and the other variants. The forward primer targets a region of the exon common to all five isoforms.
**b.** Representative RT-PCR results from SY5Y cells previously treated with anti-*CELF1* siRNA (lanes 4–6) or control siRNA (lanes 1–3). Upper gel, combination of primers F+R2. Lower gel, combination of primers F+R1. The identities of the PCR products were confirmed by sequencing, and the position of vE is indicated. Right, percentage of vE (intensity of the vE product divided by the sum of the intensities of all products). Error bars indicate the S.E.

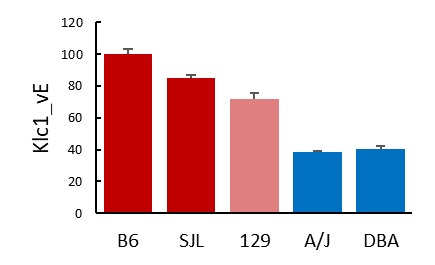

**Supplementary Fig. 4 Expression levels of *Klc1_vE* mRNA in the brains of five mouse strains at 12 months of age**

*Klc1* mRNA levels in each high amyloid-β (Aβ) strain (B6, SJL, and 129, shown with red and pink bars) were significantly higher (Tukey-Kramer HSD test, p<0.0001–0.0009) than those in each of the low Aβ strains (A/J and DBA, shown with blue bars). Error bars indicate the S.E. n=6–8 for each strain. A total of 38 animals were included.

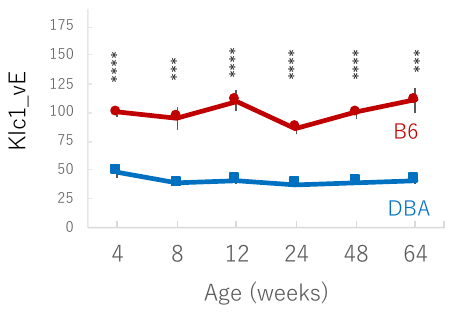

**Supplementary Fig. 5 Expression levels of *Klc1_vE* in C57BL/6 and DBA/2 mouse brains**

*Klc1_vE* mRNA levels were measured in non-Tg mouse brains at 4, 8, 12, 24, 48, and 64 weeks of age (n=5–8/group, total number of animals, 65). At all ages, the *KLC1_vE* levels were significantly lower (nominal p-value <0.0001–0.0005) in DBA/2 mice (blue) than those in C57BL/6 mice (red). Error bars indicate the S.E. *** p<0.001, **** p<0.0001.
